# Face masks to prevent transmission of COVID-19: a systematic review and meta-analysis

**DOI:** 10.1101/2020.10.16.20214171

**Authors:** Yanni Li, Mingming Liang, Liang Gao, Mubashir Ayaz Ahmed, John Patrick Uy, Ce Cheng, Qin Zhou, Chenyu Sun

## Abstract

**Background:** Based on the current status of the COVID-19 global pandemic, there is an urgent need to systematically evaluate the effectiveness of wearing masks to protect public health from COVID-19 infection.

**Methods:** We conducted a systematic review and meta-analysis to evaluate the effectiveness of using face masks to prevent the spread of SARS-CoV-2. Relevant articles were retrieved from PubMed, Web of Science, ScienceDirect, Cochrane Library, and Chinese National Knowledge Infrastructure (CNKI), VIP (Chinese) database. There were no language restrictions. This study was registered with PROSPERO under the number CRD42020211862.

**Results:** A total of 6 case-control studies were included. In general, wearing a mask was associated with a significantly reduced risk of COVID-19 infection (OR = 0.38, 95% CI = 0.21-0.69, *I*^*2*^ = 54.1%). Heterogeneity modifiers were investigated by subgroup analysis. For healthcare workers group, masks were shown to have a reduce risk of infection by nearly 70%. Studies in China showed a higher protective effect than other countries. Adjusted estimates and subgroup analyses showed similar findings.

**Conclusions:** The results of this systematic review and meta-analysis support the conclusion that wearing a mask could reduce the risk of COVID-19 infection.

## Introduction

Coronavirus disease 2019 (COVID-19) is a global pandemic that has become a major public health burden worldwide. It has many potential long-term effects due to already fragile healthcare systems(1). Severe Acute Respiratory Syndrome Coronavirus 2 (SARS-CoV-2) is transmitted through close contact and person-to-person transmission and causes COVID-19. To date, viral RNA has been found in air sampling in several studies(2, 3). For the current foreseeable future, until a safe and effective vaccine or treatment is available, COVID-19 prevention will continue to rely on non-pharmacological interventions, including mitigation of pandemics in community settings. Therefore, evaluation of personal protective equipment (PPE), such as masks or respirators, is critical to prevent the spread of SARS-CoV-2.

There are different standards of masks, and qualified masks can help to protect users from a large number of respiratory droplets(4, 5). They vary in thickness and permeability. N95 respirators are specifically designed to protect users from small airborne particles, including aerosols. Asadi et al. found that surgical masks and unventilated KN95 respirators reduced the emission rate of outward particles by an average of 90% and 74% during talking and coughing, respectively(6). In the prevention and control of COVID-19, the correct use of personal protective equipment is one of the most important measures to effectively interrupt the spread of the SARS-CoV-2, and to protect the safety of healthcare workers and other non-healthcare populations.

Recommendations regarding the effect of wearing a mask on the prevention of respiratory virus transmission (RVI) have been confirmed by many studies. A meta-analysis found reduced spread of Severe acute respiratory syndrome (SARS) (OR = 0.32; 95% CI 0.25–0.40)(7). Another meta-analysis recently found that mask use by healthcare and non-healthcare workers reduced the risk of laboratory-confirmed respiratory viral infection by 80% (95% CI = 0.11-0.37) and 47% (95% CI = 0.36-0.79), respectively(8).

Compared with other respiratory virus infections, the protective effect of masks on COVID-19 still lacks relevant comprehensive evidence. Therefore, we performed a systematic review and meta-analysis to evaluate the effectiveness of the use of masks to prevent SARS-CoV-2 transmission.

## Methods

### Identification and selection of studies

The Preferred Reporting Items for Systematic Reviews and Meta-Analysis (PRISMA) statement was consulted to report this systematic review. We prospectively submitted the systematic review protocol for registration on PROSPERO (CRD42020211862).

Regarding this meta-analysis, a comprehensive searching strategy was carefully designed to select eligible studies from multiple electronic databases, including PubMed, Web of Science, Cochrane Library, and Chinese National Knowledge Infrastructure (CNKI), VIP (Chinese) database. All included studies were published before October 2020. The following combined search terms were used in the search: (“mask” OR “face mask” OR “respirators” OR “N95” OR “*mask”) AND (“severe acute respiratory syndrome coronavirus 2” OR “2019-nCoV” OR “COVID-19” OR “SARS-CoV-2”). Relevant Chinese technical terms for the Chinese databases were used to search for published articles.

Furthermore, references of all relevant articles and reviews were retrieved to search for additional eligible studies. Articles providing abstracts only were excluded. After deleting duplicates, all abstracts and titles were filtered independently by two reviewers to remove the irrelevant articles. We downloaded and read the full text of the potential research related to the selection criteria to incorporate systematic reviews. Reviewers compared and discussed the results. If a discussion by the two reviewers did not result in an agreement, then the third party was called upon to create consensus.

### Inclusion and exclusion criteria

The studies meeting the following criteria were included: (1) concerning the relationship between the face mask and preventing COVID-19; (2) diagnosis of SARS-CoV-2 must have laboratory evidence; (3) providing complete data of cases and controls for calculating an odds ratio (OR) with 95% confidence interval (CI); (4) study design is correct and appropriate; (5) no language restrictions applied. The exclusive criteria were as follows: (1) insufficient data to ascertain the adjusted ORs; (2) conferences/meetings abstracts, case reports, editorials, and review articles; (3) duplicate publication or overlapping studies.

### Study quality assessment

The Newcastle-Ottawa Scale (NOS) was used to evaluate the quality of the case-control study: study ratings of seven to nine stars corresponded to high-quality, five to six stars to moderate quality, and four stars or less to low quality (9). Three members of the review team completed assessments independently. The disagreements were resolved by discussion.

### Statistical analysis

The association of mask use with subsequent COVID-19 was assessed with odds ratios (ORs) with a 95% confidence interval (CI). Adjusted and unadjusted pooled estimates were calculated separately. *P* values less than 0.05 were considered statistically significant. Considering the potential for between-study heterogeneity, subgroup analyzes were carried out based on stratification by HCWs, countries, and types of masks. Sensitivity analysis was performed by omitting individual studies to assess the stability of the meta-analysis. The heterogeneity was assessed using the *I*^*2*^ statistic. The heterogeneity was considered insignificance when *P* > 0.10 and *I*^*2*^ < 50%. If the study lacked heterogeneity, the pooled OR estimate was calculated using the fixed-effects model, otherwise the random-effects model was used(10). Begg’s and Egger’s test were performed to quantitatively analyze the potential publication bias. The *P* values of Begg’s and Egger’s test more than 0.05 implied no obvious publication bias in this meta-analysis(11, 12). The meta-analysis was performed using by Stata (version 14.0; Stata Corp, College Station, TX) software.

## Results

### Characteristics of eligible studies

A flow diagram of the literature search and related screening process is shown in Figure 1. A total of 6 studies met our inclusion criteria(13-18), all studies included were case-control studies (**Table 1**). Among them, studies were conducted in China, the USA, Thailand, and Bangladesh. All patients had laboratory evidence. The study by Doung-ngern et al. investigated non-professional populations, and other studies focused on healthcare workers.

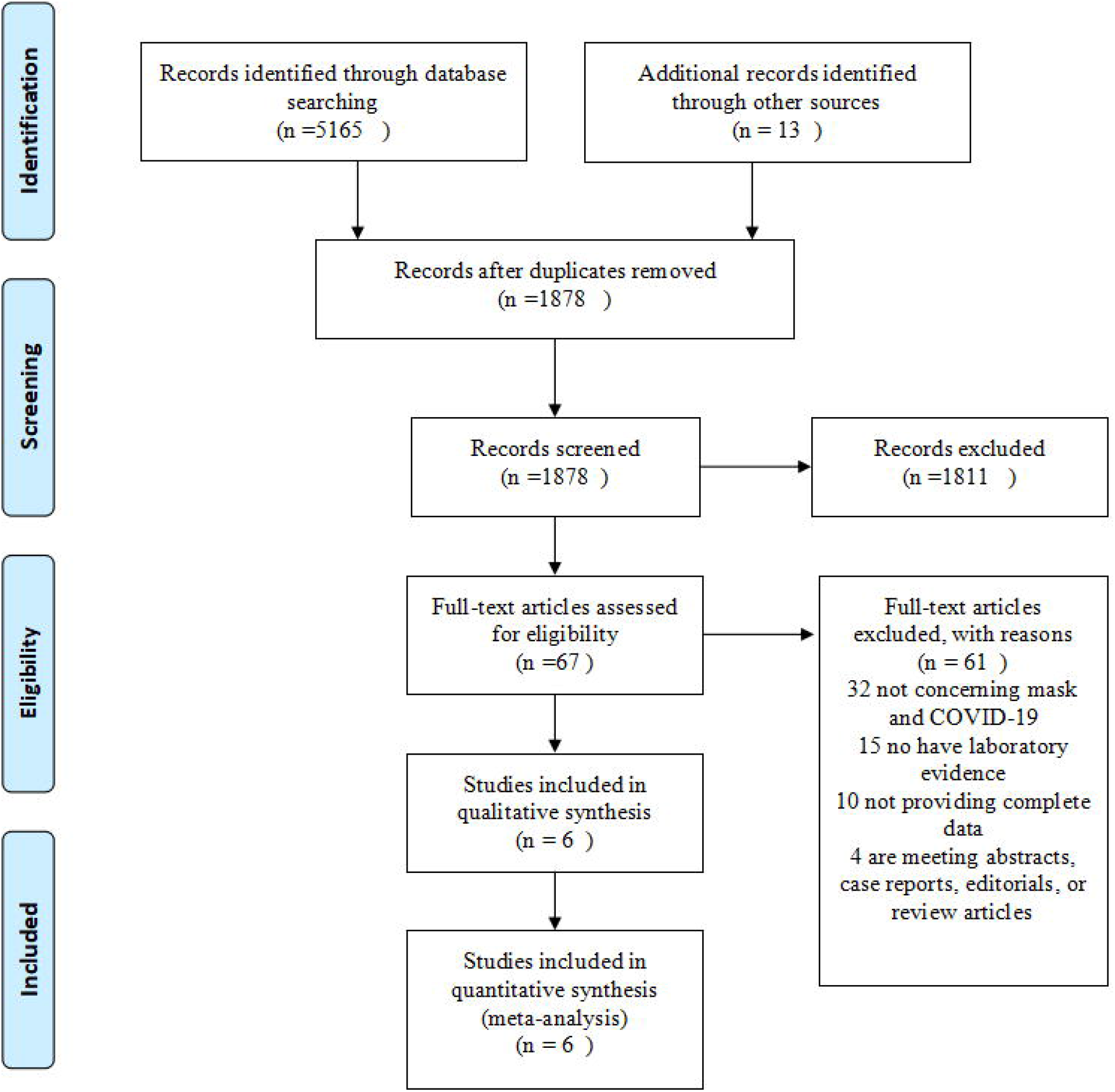

### Quality of studies

Inter-rater agreement of the quality of included studies was strong. Table 2 summarizes the quality evaluations of the included studies. Funnel plots assessing the risk of publication bias are included in figure 2. Neither Begg’s test (z=0.75, p=0.452) nor Egger’s test (t=-1.44, p=0.224) manifested any distinct evidence of the publication bias. The sensitivity analyses did not substantially alter the pooled ORs by excluding one-by-one study, indicating that the meta-analysis was generally robust.

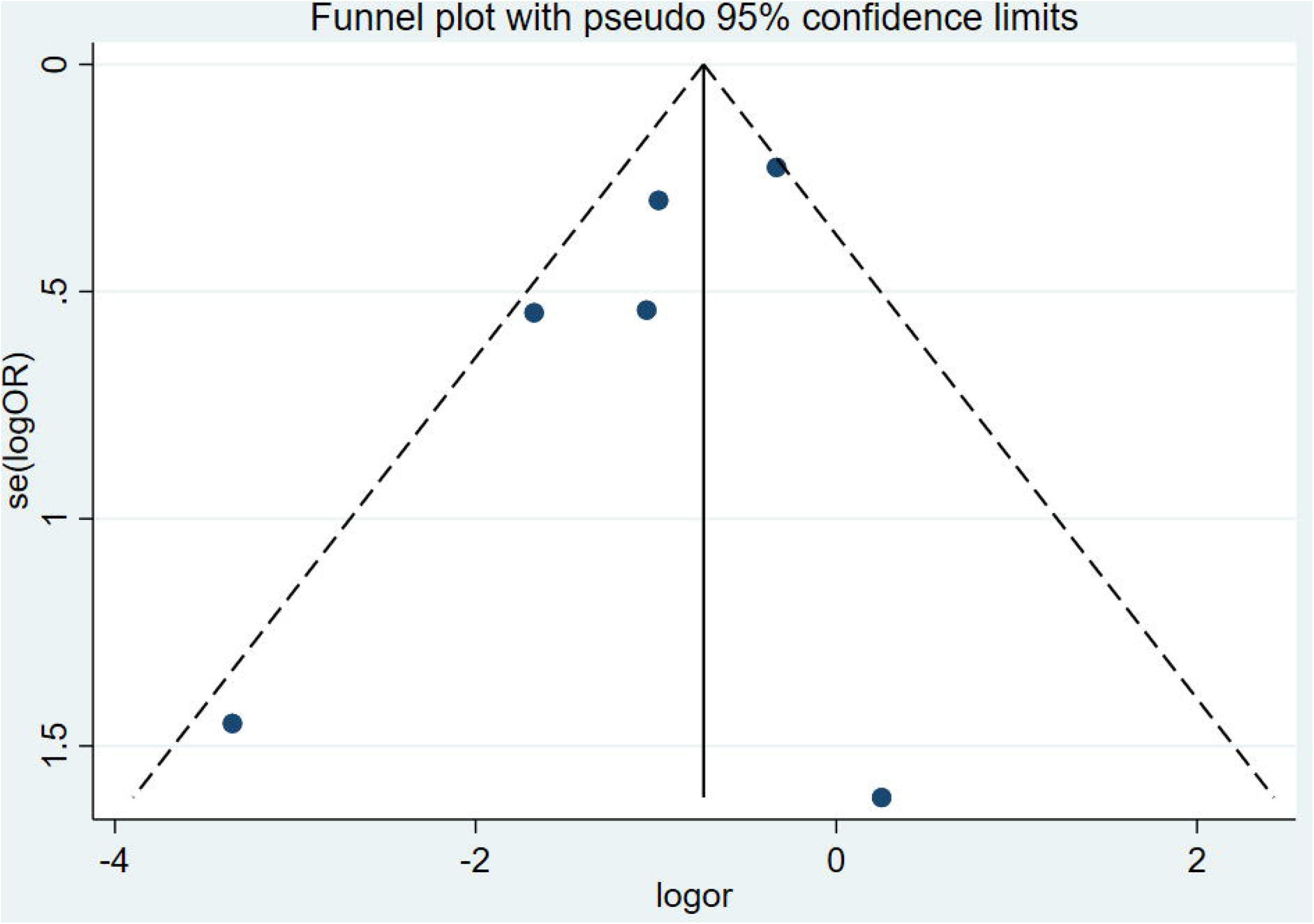

### Masks and risk of SARS-CoV-2 infection

The 6 studies reporting on the effectiveness of wearing masks included 1,233 participants. In general, face masks were effective in preventing the spread of SARS-CoV-2. After wearing a mask, the risk of contracting COVID-19 was significantly reduced, with the pooled OR of 0.38 and 95% CI=0.21-0.69 (*I*^*2*^=54.1%, M-H Random-effect model) (Figure 3).

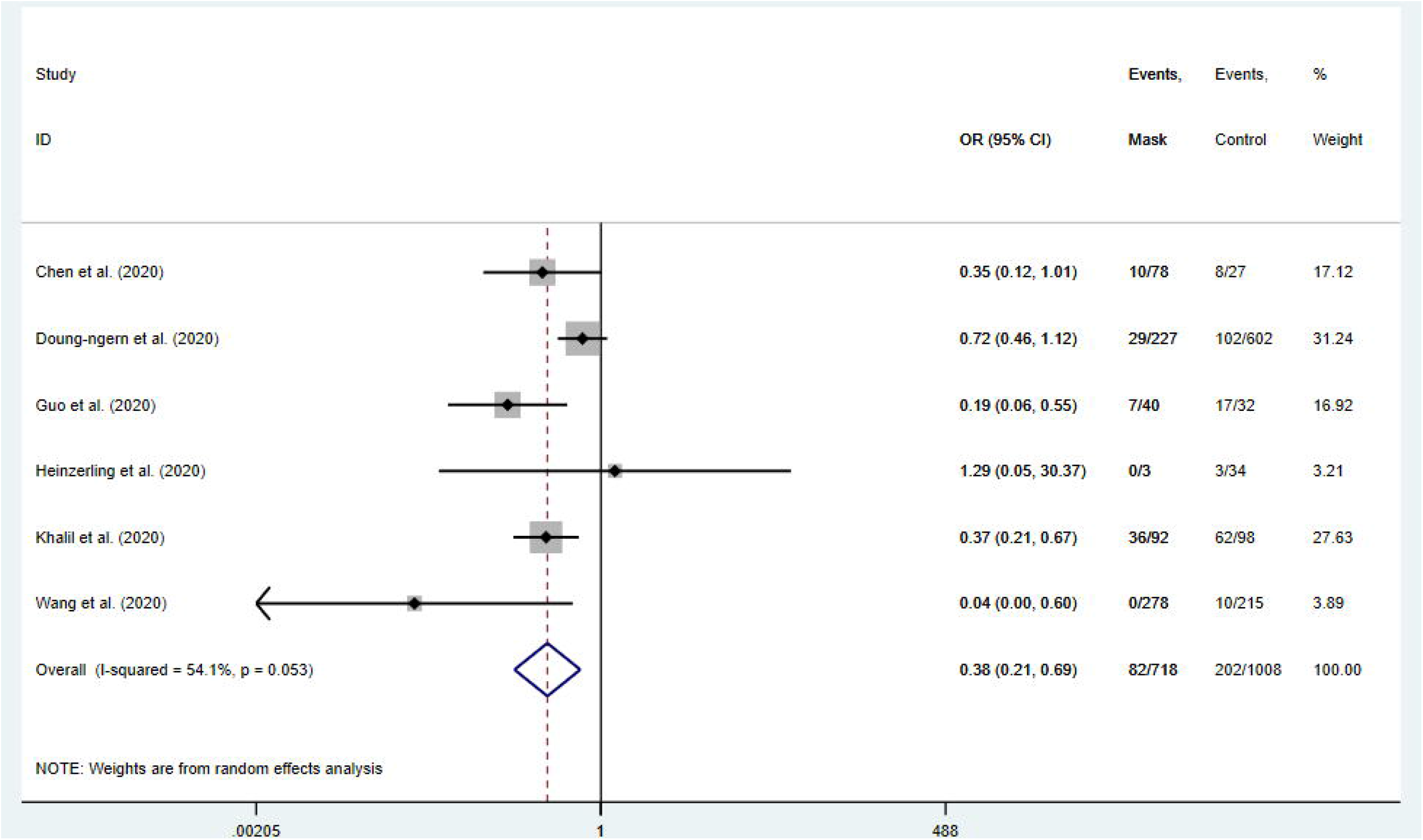

In the subgroup of HCWs only, the protective effect was more obvious, with the pooled OR of 0.29 (95% CI=0.18-0.44, *I*^*2*^=11%) (Figure 4).

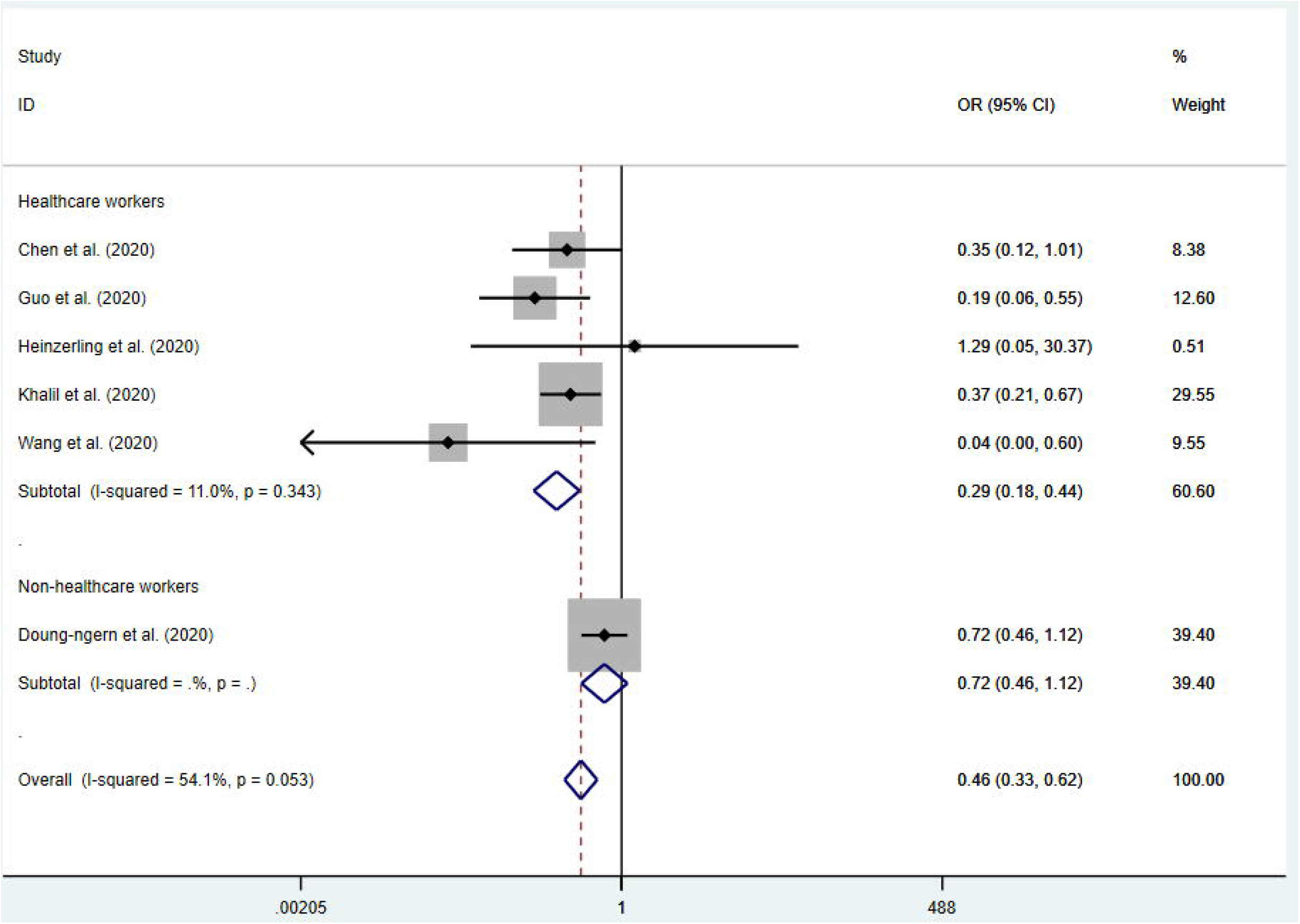

Only Doung-ngern et al. (14) investigating non-HCW, and no protective effect was found in the subgroup analysis (OR= 0.72, 95% CI=0.46-1.12). It should be noted that, the estimate after adjusted possible confounding variables was 0.23 (95% CI = 0.09-0.59) in this study, and the protection of masks was still statistically significant (adjustment variables including sex, age, contact place, shortest distance of contact, duration of contact, sharing dishes or cigarettes and handwashing) (Table 3).

By geographic locations, beneficial protective effects of wearing masks were found in China (OR=0.21, 95% CI=0.09-0.53, *I*^*2*^=26.1%), and other countries (OR=0.55, 95% CI=0.32-0.95, *I*^*2*^=39.3%). Face mask could significantly reduce the risk of SARS-CoV-2 infection (OR=0.44, 95% CI=0.21-0.93, *I*^*2*^=52.0%). And no significant protective effect was shown in N95 respirator group (OR=0.17, 95% CI=0.02-1.69, *I*^*2*^=94.6%). However, the N95 respirator showed a significant protective effect in the adjusted estimation subgroup analysis (OR=0.19, 95% CI=0.09-0.38, *I*^*2*^=0.0%) (Table 3).

## Discussion

This meta-analysis of all available articles provides the most current evidence to date on the efficacy of face masks in preventing the transmission of SARS-CoV-2, which causes COVID-19 in 2019. It spread quickly after being discovered from Wuhan, China at the end of 2019, eventually leading to a global pandemic(19). Experimental studies have grown live viruses from aerosols and surfaces several hours after implantation(20). A large amount of pathological evidence shows that aerosol transmission is the predominate route of transmission(21). Proximity and ventilation are also determinants of transmission risk(22).

Wearing a mask can prevent the inhalation of large droplets and sprays(23). Research evidence shows that masks can filter sub-micron dust particles(24). The previous meta-analysis concluded that after wearing a mask, the risk of respiratory viral infections including influenza, SARS, and H1N1 was significantly reduced with the pooled OR was of 0.35. This result is similar to the result of our meta-analysis that wearing a mask is also very effective in preventing the spread of COVID-19 (OR=0.38, 95% CI=0. 21-0.69).

The WHO started recommending wearing masks as part of a comprehensive approach to reducing the spread of SARS-CoV-2 in June, 2020(25). This is consistent with the recommendations made by the Chinese health department at the beginning of the epidemic(26). In our results, the use of face masks reduced the risk of COVID-19 infection by 70% for health care workers. However, we only included one study on the general population. In this study, the adjusted OR value was also statistically significant. Besides s, a cohort study in Beijing found that the use of masks in index patients was independently associated with a reduction in the risk of household infection(27). Investigation of the outbreak on USS Theodore Roosevelt found that low infection risk was related to self-reports of face coverings and wearing masks(28). This evidence suggests the protective effect of masks on the general population.

The United States Centers for Disease Control and Prevention specifies that the mask recommendation should not include medical masks(29). Because these masks should be reserved for healthcare workers. Regarding the types of masks, both N95 masks and general masks have been found effective in this study. This is consistent with the conclusions of previous studies. The Cochrane system review of Jefferson et al. showed that both N95 and surgical masks can effectively prevent the spread of respiratory viruses(7). And Long et al. indicated a protective effect of N95 respirators against laboratory-confirmed bacterial colonization (RR = 0.58, 95% CI 0.43-0.78)(8). However, the current available evidence has not yet confirmed the difference in protective effectiveness between N95 masks and medical surgical masks (30), although when tested in the laboratory, it was found that N95 respirators were generally more effective than surgical masks and have better facial sealing characteristics(7, 31). As face seal is critical for the N95 respirator to provide its protective effect at maximal capacity, improperly donning and doffing, or adjusting of the N95 respirator could lead to inadvertent contamination and air leak around the edge of N95 respirator, thus negating the potential protective benefit(7, 31).

This investigation also had several limitations. First, all the included studies were case-control studies and lacked adequately designed and high-quality randomized controlled studies. This may reduce the overall strength of the results. Second, because more research is currently focused on the treatment and pathology of COVID-19, the total sample size of studies on the effectiveness of PPE is still relatively small. We will continue to focus on the progress of relevant population-based studies. Third, the available studies that provided data for some subgroup analyses were limited, thus the statistical power was relatively low and the results should be interpreted with caution. Fourth, this study performed meta-analysis on the unadjusted and adjusted data to calculate the corresponding results, however, the included original studies did not make the same adjustments for possible confounding factors, such as gender, age, vaccination, hand hygiene, age, gender and cultural difference, and thus, the heterogeneity of the final results may be affected.

## Conclusion

This meta-analysis aims to provide comprehensive evidence to identify the risk of SARS-CoV-2 infection associated with mask wearing. This could help healthcare workers, public health professionals, and policy makers to identify risk factors and develop strategies to reduce COVID-19 infection. The results show that the mask has a significant protective effect against COVID-19. However, more evidence is still needed to better define the protective effect of the mask on the wider population, and more large practical trials are needed to evaluate the efficacy of the mask on the face to prevent transmission of SARS-CoV-2.

## Data Availability

Some or all data, models, or code generated or used during the study are available from the corresponding author by request.

## Acknowledgments

This work not received any funding.

## CRediT statements

*Mingming Liang &Yanni Li:* Data curation, Writing, Original draft preparation, Software. *Liang Gao:* Data curation, Conceptualization, Methodology. *John Patrick Uy:* Data curation and Writing-Reviewing. *Mubashir Ayaz Ahmed:* Data curation, Visualization. *Ce Cheng:* Data curation, Visualization. *Qin Zhou:* Data curation, Visualization. *Chenyu Sun:* Supervision, Writing-Reviewing and Editing.

All authors read and approved the final manuscript.

