## Supplementary material for "Face masks to prevent transmission of COVID-19: a systematic review and meta-analysis": table 1

Table 1 Characteristics of eligible studies

|  | Study | Year | Country | Virus | Infections | Mask group | Infections | | Control group | Mask type | Type of Study | Healthcare workers | Main findings & comments |
| --- | --- | --- | --- | --- | --- | --- | --- | --- | --- | --- | --- | --- | --- |
| 1 | Chen et al. | 2021 | China | 2019-nCoV | 10 | 78 | 8 | 27 | | Mask^*^ | Case-control study | Healthcare workers | Risk analysis revealed that wearing face masks could reduce infection risk. |
| 2 | Doung-ngern et al. | 2020 | Thailand | 2019-nCoV | 29 | 227 | 102 | 602 | | Mask | Case-control study | Non-healthcare workers | The consistent wearing of masks, handwashing, and social distancing to protect against COVID-19. |
| 3 | Guo et al. | 2020 | China | 2019-nCoV | 7 | 40 | 17 | 32 | | Mask | Case-control study | Healthcare workers | Wearing respirators or masks all of the time was found to be protective. |
| 4 | Heinzerling et al. | 2020 | USA | 2019-nCoV | 0 | 3 | 3 | 34 | | Mask | Case-control study | Healthcare workers | PPE use can help minimize unprotected, high-risk HCP exposures and protect the health care workforce. |
| 5 | Khalil et al. | 2020 | Bangladesh | 2019-nCoV | 36 | 92 | 62 | 98 | | N95 | Case-control study | Healthcare workers | The use of masks and decontamination of the patient’s surroundings may give protection against COVID-19. |
| 6 | Wang et al. | 2020 | China | 2019-nCoV | 0 | 278 | 10 | 215 | | N95 | Case-control study | Healthcare workers | The 2019-nCoV infection rate for medical staff was significantly increased in the no-mask group compared with the N95 respirator group (adjusted odds ratio (OR): 464.82, [95% CI: 97.73-infinite] ). |

* Specific type of mask was not reported
