## Supplementary material for "Face masks to prevent transmission of COVID-19: a systematic review and meta-analysis": table 2

Table 2 The quality of the case-control studies and cohort studies

|  | Study | Year | Selection | Comparability | Outcome | Stars* |
| --- | --- | --- | --- | --- | --- | --- |
| 1 | Gan | 2020 | 4 | 2 | 2 | 8 |
| 2 | Mohammad | 2018 | 4 | 2 | 2 | 8 |
| 3 | Gebhard | 2018 | 3 | 1 | 2 | 6 |
| 4 | Auger | 2017 | 3 | 2 | 2 | 7 |
| 5 | Hopstock | 2012 | 3 | 2 | 2 | 7 |
| 6 | Southern | 2006 | 3 | 1 | 2 | 6 |
| 7 | Gerber | 2006 | 4 | 2 | 2 | 8 |
| 8 | Ohlson | 1991 | 3 | 1 | 2 | 6 |

* Scoring by Newcastle-Ottawa Scale
