## Supplementary material for "Face masks to prevent transmission of COVID-19: a systematic review and meta-analysis": table 3

Table 3. Meta-analysis results of the effect of masks on COVID-19 infection among different subgroups.

|  | Subgroup | Study numbers | OR | 95%CI | Heterogeneity |
| --- | --- | --- | --- | --- | --- |
| Unadjusted estimates | Overall | 6 | 0.38 | 0.21-0.69 | 54.1% |
|  | HCWs | 5 | 0.29 | 0.18-0.44 | 11.0% |
|  | Non-HCWs | 1 | 0.72 | 0.46-1.12 | N/A |
|  | China | 3 | 0.21 | 0.09-0.53 | 26.1% |
|  | Other countries | 3 | 0.55 | 0.32-0.95 | 39.3% |
|  | Mask group* | 4 | 0.44 | 0.21-0.93 | 52.0% |
|  | N95 group | 2 | 0.17 | 0.02-1.69 | 64.6% |
| Adjusted estimates | Overall | 5 | 0.19 | 0.11-0.33 | 77.6% |
|  | HCWs | 4 | 0.18 | 0.09-0.34 | 83.0% |
|  | Non-HCWs | 1 | 0.23 | 0.09-0.59 | N/A |
|  | China | 3 | 0.06 | 0.02-0.17 | 81.4% |
|  | Other countries | 2 | 0.3 | 0.16-0.57 | 0.0% |
|  | Mask group | 3 | 0.19 | 0.09-0.38 | 0.0% |
|  | N95 group | 2 | 0.2 | 0.09-0.44 | 94.3% |

HCW: Healthcare workers; Non-HCWs: Non-healthcare workers; N/A: Not applicable; * Specific type of mask was not reported
